# Age-related alterations in blood fatty acid composition and ratios, omega-3 fatty acid Levels and association with PhenoAge acceleration: a cross-sectional analysis of the NHANES 2021-2023 data

**DOI:** 10.64898/2026.09.21.26363464

**Authors:** Anne Pietzner, Nadine Rohwer, Karsten H. Weylandt

## Abstract

**Introduction:** Polyunsaturated fatty acid (PUFA) status is an important marker of metabolic health across the lifespan, and omega-3 (n-3) PUFA were shown to have anti-inflammatory and cardioprotective effects. The omega-3 index, a composite of eicosapentaenoic acid (EPA) and docosahexaenoic acid (DHA), is widely used to assess n-3 PUFA supplementation status, and product-to-precursor fatty acid ratios are used as proxy indices of desaturation and elongation. Previous data indicate a higher proportion of n-3 PUFA with age. Whether the same pattern can be found in an NHANES cohort, and whether long-chain n-3 status relates to phenotypic age (PhenoAge), a composite measure of biological age, has not been established.

**Methods:** We analyzed erythrocyte fatty acid profiles measured by electron capture negative-ion mass spectrometry in erythrocytes of participants of the National Health and Nutrition Examination Survey (NHANES) 2021-2023. Adults aged 20-79 years with a complete 21-fatty-acid panel were included (n = 4,863) and stratified into five age groups (20-34, 35-44, 45-54, 55-64, 65-79 years). In the fasting subsample with a measured fasting glucose (n = 2,822) we additionally computed PhenoAge according to Levine and assessed PhenoAge acceleration (PhenoAge regressed on chronological age) in relation to EPA and DHA.

**Results:** Percentage of summarized PUFA and n-6 PUFA decreased with age while percentage of n-3 PUFA and EPA + DHA increased. In contrast, total saturated fatty acids did not differ across age groups. EPA and DHA rose with age, whereas linoleic acid (LA), dihomo-γ-linolenic acid (DGLA) and α-linolenic acid (ALA) declined. The delta-5-desaturase (D5D) index (AA/DGLA) and the AA/LA ratio were positively associated with age, while elongation-related proxies ELOVL2 (DTA/AA, DPA/EPA) and ELOVL5 (DGLA/GLA) were inversely associated.

Higher EPA + DHA was associated with lower PhenoAge acceleration with the most pronounced effect of - 4,7 years in the group with highest (≥8%) EPA+DHA content.

**Discussion:** Age-related shifts in erythrocyte PUFA composition and in desaturase and elongase proxy indices reported previously in a clinical cohort from Germany are reproduced in a large US population. In addition, long-chain n-3 status was inversely associated with PhenoAge acceleration.

## 1 Introduction

Omega-3 polyunsaturated fatty acids (n-3 PUFA) have been topic of intensive research since seminal studies in the 1970s implicating them in the protection from cardiovascular disease (Dyerberg et al., 1978; Dyerberg and Bang, 1979). Since then, recommendations to increase n-3 PUFA uptake have been widely discussed and established (Siscovick et al., 2017) and omega-3 (n-3) supplements are widely used supplements (Clarke et al., 2015; Kantor et al., 2016; Li et al., 2020). Uptake of n-3 PUFA has increased in recent decades in many parts of the world (Micha et al., 2014; Schuchardt et al., 2024).

However, large clinical intervention studies using pharma grade n-3 PUFA had mixed results with some positive (Yokoyama et al., 2007; Bhatt et al., 2019; Lok et al., 2026) but also some negative findings (Group et al., 2018; Manson et al., 2019; Nicholls et al., 2020).

Interestingly, analyses from a randomized controlled intervention trial indicated that n-3 supplementation modestly slowed DNA-methylation-based biological aging in older adults (Bischoff-Ferrari et al., 2025). This positions the n-3 PUFA now in the emerging field of longevity medicine, where - should these data be confirmed - the n-3 PUFAs could be a low-risk intervention associated with a gain of lifetime.

In accordance with this, large analyses indicate higher n-3 PUFA to be associated with decreased premature mortality (Harris et al., 2018; Harris et al., 2021). A cohort study based on a subset of UK Biobank participants recently demonstrated that higher levels of n-3 PUFA and of the omega-6 (n-6) PUFA linolenic acid (LA) was associated with a higher likelihood of healthy aging (Lyu et al., 2026).

We recently described in 1277 patients from a metabolic disease clinic in rural Brandenburg, Germany that participants aged ≥ 65 years had higher total n-3 and lower total n-6 PUFA levels. Eicosapentaenoic acid (EPA) and docosahexaenoic acid (DHA) increased with age while LA and dihomo-γ-linolenic acid (DGLA) decreased, and delta-5-desaturase (D5D) and arachidonic acid (AA)/LA rose with age whereas elongation of very long chain fatty acids (ELOVL)2- and ELOVL6-related ratios decreased (Yu et al., 2026).

Several previous studies have indicated that blood PUFA content and ratios may shift with age with increased levels of n-3 PUFA in older patients (Crowe et al., 2008; Harris et al., 2021; Diffenderfer et al., 2022). As aging is accompanied by progressive changes in lipid metabolism that may influence cellular homeostasis and susceptibility to age-related disease (Hornburg et al., 2023) and PUFA are essential constituents of cell membranes and precursors of bioactive lipid mediators, and the balance between the n-6 and n-3 families affects membrane properties, receptor signaling and inflammatory tone (Schmitz and Ecker, 2008; Harayama and Shimizu, 2020), these differences in circulating PUFA status could therefore have also functional consequences.

In the context of aging, the ELOVL2 locus is of particular interest: DNA methylation at the *ELOVL2* locus is a well described epigenetic marker of chronological age (Garagnani et al., 2012; Paparazzo et al., 2023).

Recently, data from the National Health and Nutrition Examination Survey (NHANES) cycle covering August 2021 to August 2023 were published, being the first to report erythrocyte fatty acid composition for a probability sample of the US population (Powers et al., 2026). This dataset also shows an increase in n-3 PUFA with age, although a thorough analysis of age-related changes in fatty acids has to our knowledge not been performed in this dataset. The NHANES 2021-2023 data therefore permit a direct assessment of some of the observations made and hypotheses generated previously.

The NHANES 2021-2023 data also allow further analyses of routine blood parameters and thus also offer the possibility to analyze biomarkers needed to calculate the so-called phenotypic age (PhenoAge), a measure of biological age developed from previous NHANES data cycles: PhenoAge is calculated from a composite of chronological age and nine routine clinical laboratory markers and predicts mortality risk (Levine et al., 2018; Liu et al., 2018); PhenoAge acceleration, defined as the residual of PhenoAge regressed on chronological age, is widely used as a summary measure of biological aging (Levine et al., 2018; Liu et al., 2018).

We therefore analyzed the NHANES August 2021-August 2023 erythrocyte fatty acid data with two aims: to assess the age-related PUFA and enzyme proxy-index patterns described in our previous study based on our measurements, and to test the association between the omega-3 index and PhenoAge acceleration in the same participants.

## 2 Materials and methods

Data were drawn from the NHANES cycle covering August 2021 to August 2023, a cross-sectional, multistage probability sample of the civilian non-institutionalized US population conducted by the National Center for Health Statistics (NCHS). All data files were downloaded from the public NHANES repository. NHANES is approved by the NCHS Research Ethics Review Board and all participants provided written informed consent; because the data are publicly available and de-identified, no additional ethical approval was required for the present analysis. In this NHANES cycle fatty acid analyses were performed in addition to other lab tests (US Centers for Disease Control and Prevention, 2025).

### Analysis populations

Two overlapping but non-identical samples were analyzed. The chronological-age analysis (Part A) included 4863 participants with a complete erythrocyte fatty acid profile. The biological-age analysis (Part B) included 2822 participants for whom erythrocyte EPA+DHA and all nine biomarkers required to compute Phenotypic Age were available. Of the combined sample, 2574 participants contributed to both analyses, 2289 to Part A only, and 248 to Part B only. Because the two parts address different questions and were defined by different data-availability requirements, they are reported separately and no participant-level linkage between the two sets of results is implied.

### Erythrocyte fatty acids and estimated desaturase/elongase activities across chronological age

Erythrocyte fatty acids were expressed as a percentage of total identified fatty acids. Analyses covered fatty acid classes: summarized (Σ) saturated fatty acids (ΣSFA), monounsaturated fatty acids (ΣMUFA), ΣPUFA, Σn-3, Σn-6, and EPA+DHA, individual fatty acids (alpha-linolenic acid (ALA), EPA, DHA, LA, DGLA, AA), and product-to-precursor ratios used as proxies for desaturase and elongase activities (D5D, delta-6 desaturase (D6D), stearoyl CoA-desaturase (SCD)-16, SCD-18, ELOVL2, ELOVL5, ELOVL6). Age was analyzed both categorically and continuously. For the categorical analysis, participants were assigned a priori to five age groups (20-34, 35-44, 45-54, 55-64, and 65-79 years). Differences across groups were tested with the Kruskal-Wallis test followed by Dunn’s post hoc test, using the oldest group (65-79 years) as the reference. For the continuous analysis, associations with chronological age were quantified using Spearman rank correlation (ρ), and simple linear regression was used to estimate the direction and magnitude of the trend (slope, R^2^).

### Derivation of Phenotypic Age and PhenoAge acceleration

PhenoAge was calculated from chronological age and nine clinical biomarkers (albumin, creatinine, glucose, C-reactive protein [log-transformed], lymphocyte percentage, mean corpuscular volume, red cell distribution width, alkaline phosphatase, and white blood cell count) using the published mortality-based algorithm [Liu et al. 2018; corrected equation in Liu et al. 2019], with all biomarkers converted to the units specified for that algorithm prior to calculation. PhenoAge acceleration (PhenoAgeAccel) was defined as the residual from an ordinary least-squares regression of PhenoAge on chronological age, estimated within the Part B population.

Positive values indicate biological aging that is accelerated relative to chronological age, and negative values indicate decelerated aging. By construction, PhenoAgeAccel has a mean of zero and is statistically independent of chronological age, so associations with PhenoAgeAccel are inherently adjusted for chronological age.

### EPA+DHA and PhenoAge acceleration

The relationship between erythrocyte EPA+DHA and PhenoAgeAccel was examined categorically and continuously. For the categorical analysis, participants were assigned to four predefined categories based on established omega-3 index thresholds (< 4%, 4-< 6%, 6-< 8%, and ≥ 8%). PhenoAgeAccel was compared across categories using the Kruskal-Wallis test with Dunn’s post hoc test, using the < 4% category (highest-risk range) as the reference; the global Kruskal-Wallis *p*-value is reported alongside the pairwise comparisons. For descriptive purposes, the proportion of participants with negative PhenoAgeAccel (biologically younger than expected) was calculated within each category. For the continuous analysis, the association between EPA+DHA and PhenoAgeAccel across the full Part B sample was quantified using Spearman rank correlation (ρ). A simple linear regression line is shown in the corresponding scatter plot for visualization only.

All analyses were performed in GraphPad Prism 9 [Version 9.5.1], with data preparation and derivation of variables carried out in Microsoft Excel. All tests were two-sided, and *p* < 0.05 was considered statistically significant. Because the key continuous variables were non-normally distributed (Shapiro-Wilk test), non-parametric methods were used throughout, and continuous variables are reported as median (interquartile range, IQR) unless stated otherwise. No imputation of missing data was performed; each analysis was based on participants with complete data for the respective variables.

## 3 Results

### 3.1 Study population

The analytic sample comprised 4863 adults aged 20-79 years (median 56 years [38-66]), of whom 2,688 (55.3 %) were women. Group sizes ranged from 643 in the 45-54-year stratum to 1432 in the 65-79-year stratum (Table 1). The proportion of women was similar across strata (53.9 %-57.5 %).

**Table 1.** Characteristics of the study population by age group.

| Age group (n) | Median Age, years [interquartile range] | Female |
| --- | --- | --- |
| 20-34 (n = 926) | 29 [24-32] | 516 (55.7 %) |
| 35-44 (n = 756) | 37 [35-40] | 435 (57.5 %) |
| 45-54 (n = 643) | 50 [47-52] | 369 (57.4 %) |
| 55-64 (n = 1106) | 60 [58-62] | 596 (53.9 %) |
| 65-79 (n = 1432) | 70 [67-74] | 722 (53.9 %) |

Participants were stratified into five age groups (20-34, 35-44, 45-54, 55-64, and 65-79 years); n denotes the number of participants per group. Age is presented as median [interquartile range, Q1-Q3] in years. Sex is given as the number (percentage) of women per group.

### 3.2 Age-related changes in erythrocyte fatty acid class distribution

ΣSFA did not differ across age groups (Kruskal-Wallis p = 0.399), whereas ΣMUFA increased and ΣPUFA decreased with advancing age (both p < 0.001) (Table 2). The change in ΣPUFA was driven entirely by the n-6 family: Σn-6 PUFA was higher in every younger stratum than in the 65-79-year reference group (p ≤ 0.002), while Σn-3 PUFA showed the opposite gradient, being lower in all four younger strata (all adjusted p < 0.001). The sum of EPA + DHA, a measure analogous to the omega-3 index, rose monotonically from a median of 3.82 % in the 20-34-year group to 4.36 % in the 65-79-year group (p < 0.001).

**Table 2.**
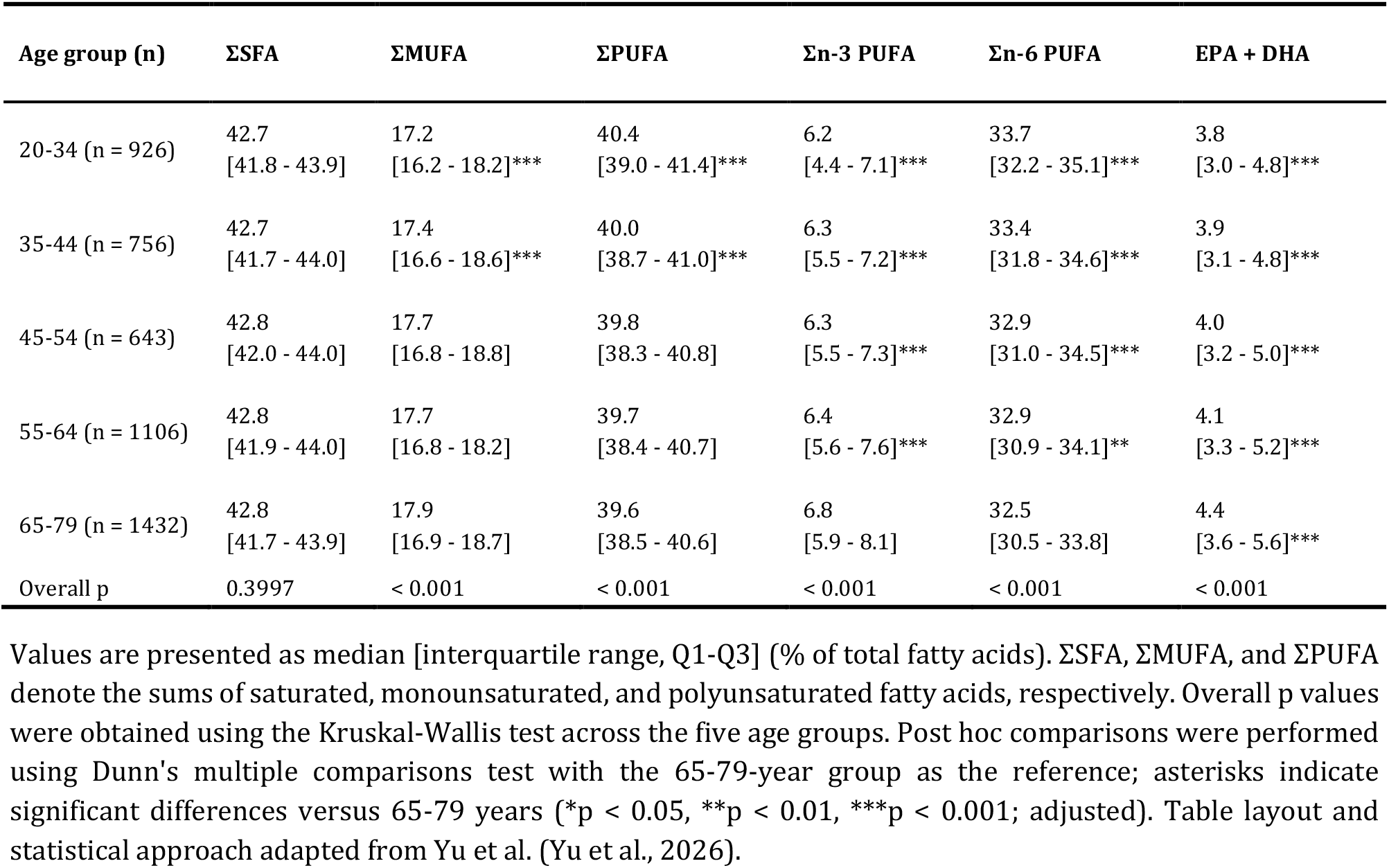
Erythrocyte fatty acid composition across age groups.

### 3.3 Individual PUFA across age groups

EPA and DHA both increased with age (Figure 1B, C). Median EPA rose from 0.4 % [0.3-0.5] in the 20-34-year group to 0.5 % [0.4-0.6] in the 65-79-year group, and median DHA from 3.5 % [2.70-4.4] to 3.9 % [3.2-5.0]; all four younger strata differed from the reference group (adjusted p < 0.05 for EPA in 55-64 years, p < 0.001 for all remaining contrasts).

**Figure 1.**
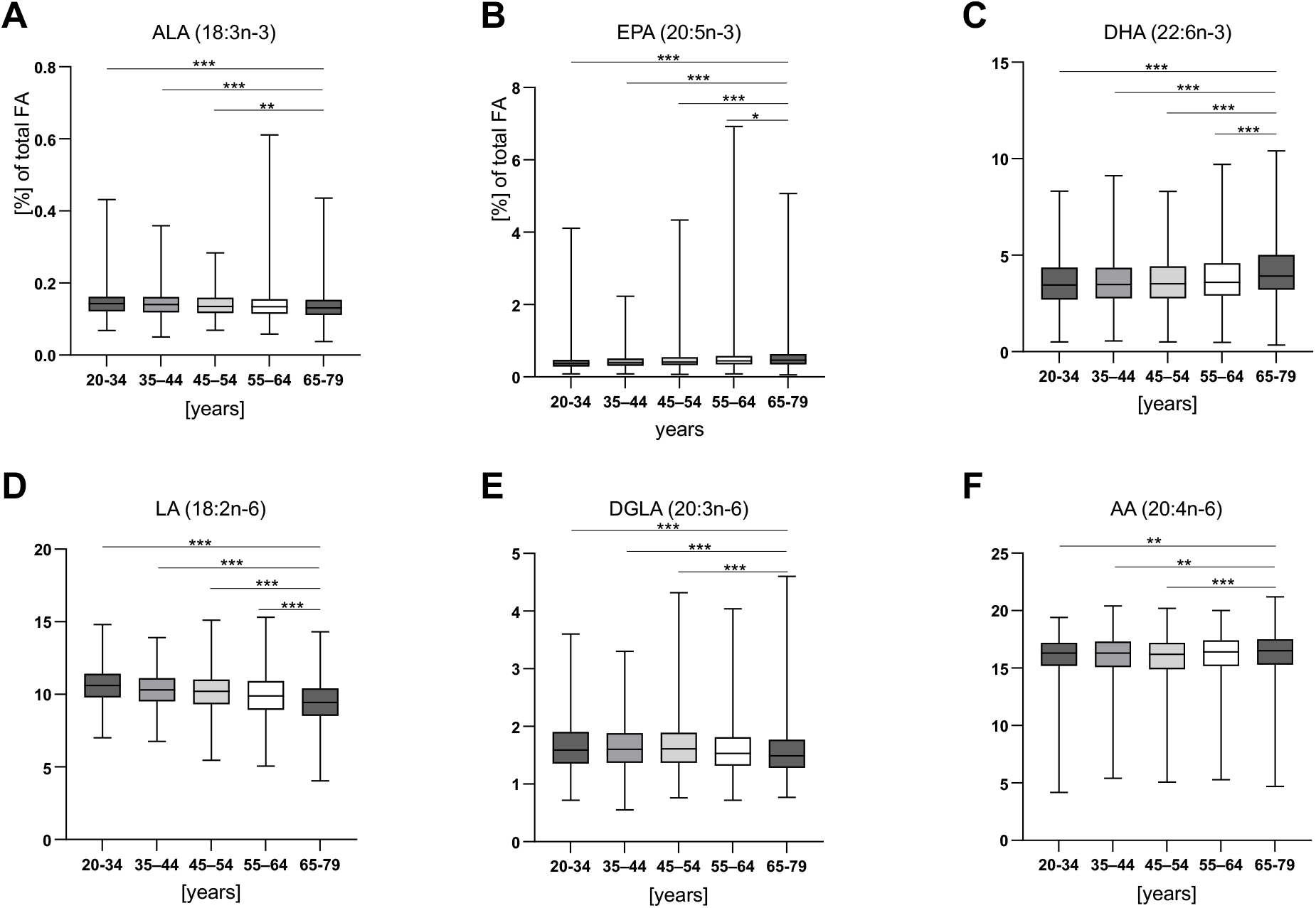
Distribution of individual n-3 and n-6 PUFA in erythrocytes across age groups. Erythrocyte levels (% of total fatty acids) are shown for the n-3 PUFA (A) α-linolenic acid (ALA, 18:3n-3), (B) eicosapentaenoic acid (EPA, 20:5n-3), and (C) docosahexaenoic acid (DHA, 22:6n-3), and for the n-6 PUFA (D) linoleic acid (LA, 18:2n-6), (E) dihomo-γ-linolenic acid (DGLA, 20:3n-6), and (F) arachidonic acid (AA, 20:4n-6). Age groups: 20-34 years (n = 926); 35-44 (n = 756); 45-54 (n = 643); 55-64 (n = 1106); 65-79 (n = 1432); total (n = 4863). Boxes show the median and interquartile range (25th-75th percentiles); whiskers extend from the minimum to the maximum value. Overall differences across the five age groups were assessed using the Kruskal-Wallis test; planned post hoc comparisons versus the 65-79-year reference group were performed using Dunn’s multiple comparisons test with multiplicity-adjusted p values. Asterisks indicate significant differences versus the 65-79-year group (*p < 0.05, **p < 0.01, ***p < 0.001; adjusted). Figure design and statistical approach adapted from Yu et al.(Yu et al., 2026).

ALA moved in the opposite direction, declining modestly from 0.143 % [0.122-0.162] in the youngest group to 0.13 % [0.11-0.15] in the oldest (overall p < 0.001; Figure 1A), with significant contrasts in the three youngest strata. This differs from the Brandenburg cohort, where erythrocyte ALA was higher in the youngest group but showed no consistent gradient across the intermediate strata (Yu et al., 2026).

LA showed the steepest and most consistent age gradient of any individual fatty acid, falling from a median of 10.6 % [9.8-11.4] in the 20-34-year group to 9.44 % [8.5-10.4] in the 65-79-year group (all contrasts adjusted p < 0.001; Figure 1D).

DGLA declined from 1.6 % [1.4-1.9] to 1.5 % [1.3-1.8] (overall p < 0.001), with significant contrasts in the three youngest strata but not in the 55-64-year group (Figure 1E).

AA showed only a small and non-monotone difference across age groups (overall p < 0.001, median 16.3 % versus 16.5 %; Figure 1F), consistent with the tighter regulation of AA observed in our previous study (Yu et al., 2026).

### 3.4 Associations of long-chain n-3 PUFA with age as a continuous variable

Treating age as a continuous variable confirmed the group-based findings (Figure 2). EPA + DHA was positively correlated with age (ρ = 0.18, p < 0.001; slope 0.017 percentage points per year, R^2^ = 0.030), as were EPA (ρ = 0.21, p < 0.001) and DHA (ρ = 0.17, p < 0.001).

**Figure 2.**
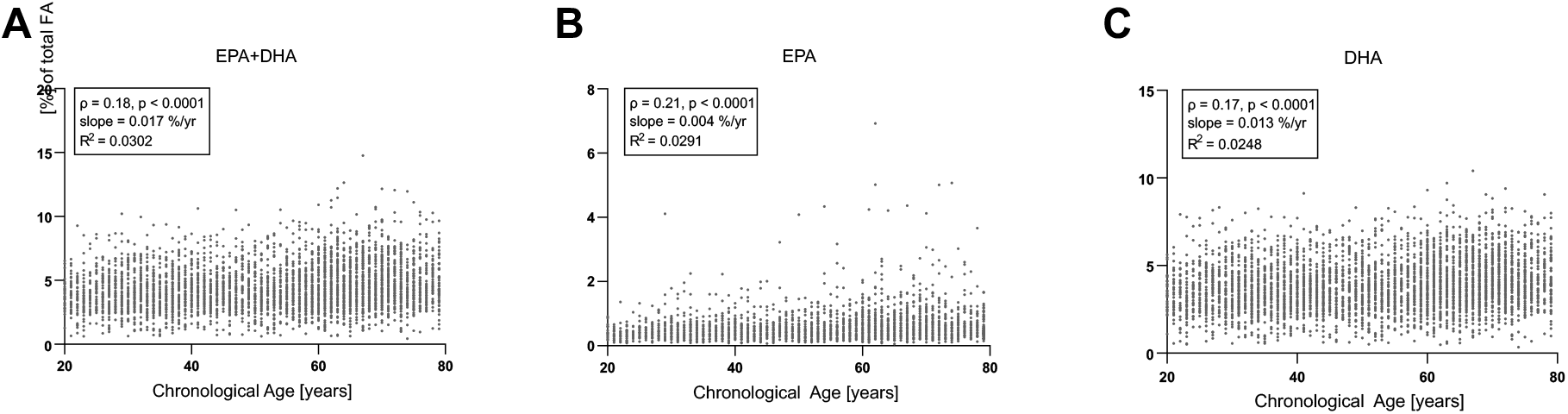
Associations of erythrocyte n-3 PUFA with age. Scatter plots show erythrocyte levels (% of total fatty acids) of (A) EPA + DHA, (B) eicosapentaenoic acid (EPA, 20:5n-3), and (C) docosahexaenoic acid (DHA, 22:6n-3) against age as a continuous variable (years). Each point represents an individual participant (total n = 4863). Solid lines denote the linear regression fit. Associations were assessed using Spearman rank correlation (ρ, two-sided p) and unadjusted linear regression with age as a continuous predictor; correlation coefficients, regression slopes, p and R^2^ are given in each panel.

### 3.5 Age-related variation in desaturase and elongase proxy indices

Ratio-based indices (Table 3) showed a similar pattern to the one we previously reported (Yu et al., 2026).

**Table 3.**
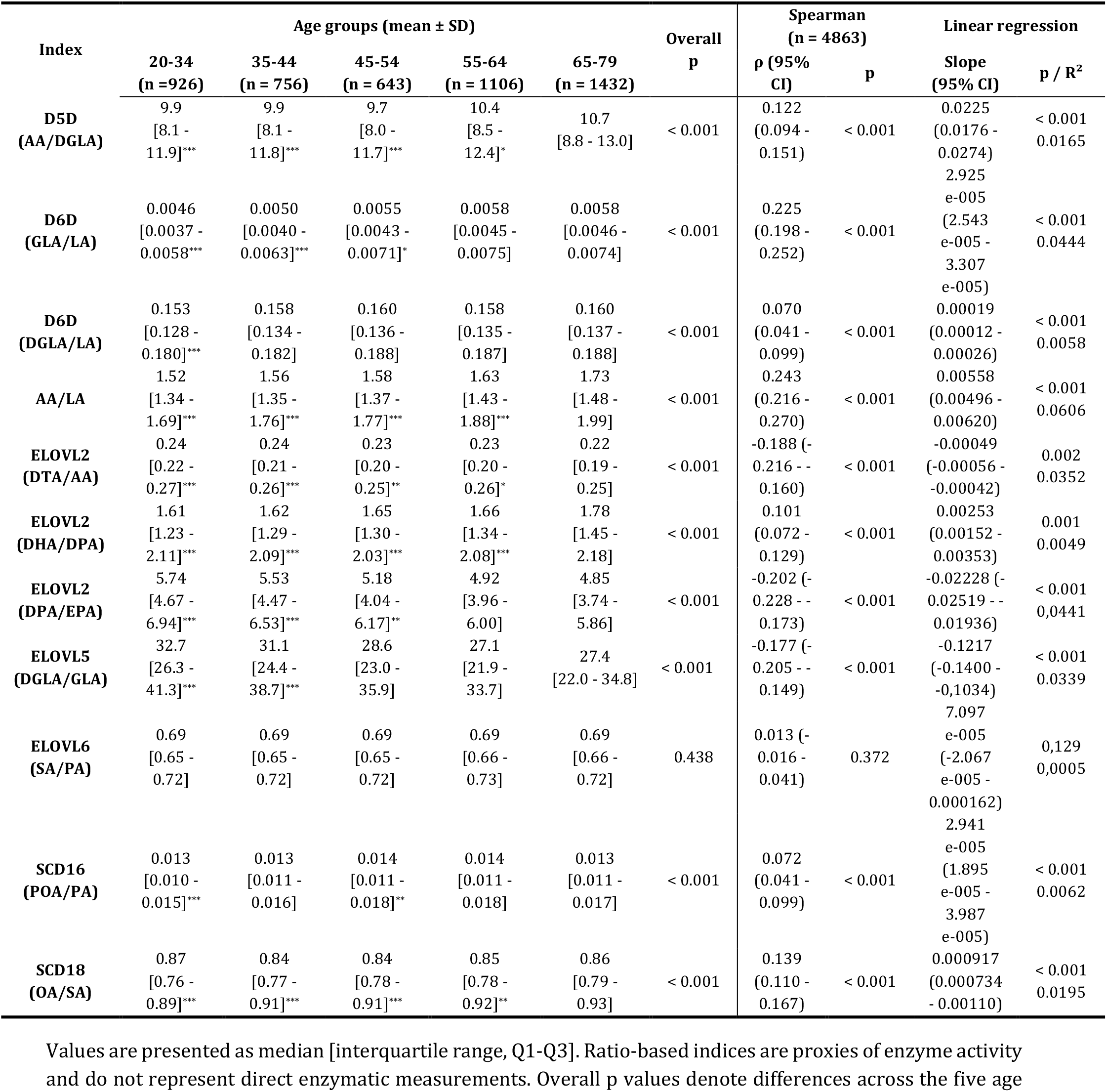
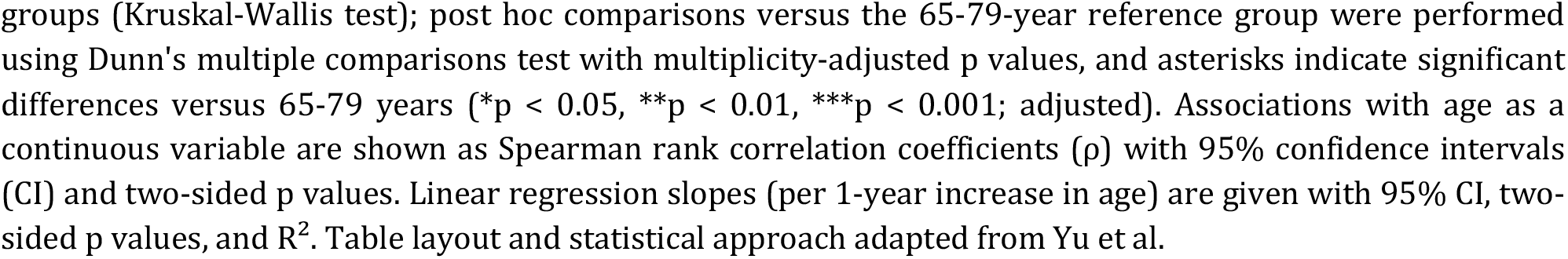
Age-stratified desaturase and elongase proxy indices and their associations with age in erythrocytes.

| Index | Age groups (mean ± SD) | | | | | Overall<br>p | Spearman<br>(n = 4863) | | Linear regression | |
| --- | --- | --- | --- | --- | --- | --- | --- | --- | --- | --- |
| | 20-34<br>(n = 926) | 35-44<br>(n = 756) | 45-54<br>(n = 643) | 55-64<br>(n = 1106) | 65-79<br>(n = 1432) | | $\rho$ (95%<br>CI) | p | Slope<br>(95% CI) | p / $R^2$ |
| **D5D**<br>**(AA/DGLA)** | 9.9<br>[8.1 -<br>11.9]*** | 9.9<br>[8.1 -<br>11.8]*** | 9.7<br>[8.0 -<br>11.7]*** | 10.4<br>[8.5 -<br>12.4]* | 10.7<br>[8.8 - 13.0] | < 0.001 | 0.122<br>(0.094 -<br>0.151) | < 0.001 | 0.0225<br>(0.0176 -<br>0.0274) | < 0.001<br>0.0165 |
| **D6D**<br>**(GLA/LA)** | 0.0046<br>[0.0037 -<br>0.0058]*** | 0.0050<br>[0.0040 -<br>0.0063]*** | 0.0055<br>[0.0043 -<br>0.0071]* | 0.0058<br>[0.0045 -<br>0.0075] | 0.0058<br>[0.0046 -<br>0.0074] | < 0.001 | 0.225<br>(0.198 -<br>0.252) | < 0.001 | 2.925<br>e-005<br>(2.543<br>e-005 -<br>3.307<br>e-005) | < 0.001<br>0.0444 |
| **D6D**<br>**(DGLA/LA)** | 0.153<br>[0.128 -<br>0.180]*** | 0.158<br>[0.134 -<br>0.182] | 0.160<br>[0.136 -<br>0.188] | 0.158<br>[0.135 -<br>0.187] | 0.160<br>[0.137 -<br>0.188] | < 0.001 | 0.070<br>(0.041 -<br>0.099) | < 0.001 | 0.00019<br>(0.00012 -<br>0.00026) | < 0.001<br>0.0058 |
| **AA/LA** | 1.52<br>[1.34 -<br>1.69]*** | 1.56<br>[1.35 -<br>1.76]*** | 1.58<br>[1.37 -<br>1.77]*** | 1.63<br>[1.43 -<br>1.88]*** | 1.73<br>[1.48 -<br>1.99] | < 0.001 | 0.243<br>(0.216 -<br>0.270) | < 0.001 | 0.00558<br>(0.00496 -<br>0.00620) | < 0.001<br>0.0606 |
| **ELOVL2**<br>**(DTA/AA)** | 0.24<br>[0.22 -<br>0.27]*** | 0.24<br>[0.21 -<br>0.26]*** | 0.23<br>[0.20 -<br>0.25]** | 0.23<br>[0.20 -<br>0.26]* | 0.22<br>[0.19 -<br>0.25] | < 0.001 | -0.188 (-<br>0.216 - -<br>0.160) | < 0.001 | -0.00049<br>(-0.00056 -<br>-0.00042) | 0.002<br>0.0352 |
| **ELOVL2**<br>**(DHA/DPA)** | 1.61<br>[1.23 -<br>2.11]*** | 1.62<br>[1.29 -<br>2.09]*** | 1.65<br>[1.30 -<br>2.03]*** | 1.66<br>[1.34 -<br>2.08]*** | 1.78<br>[1.45 -<br>2.18] | < 0.001 | 0.101<br>(0.072 -<br>0.129) | < 0.001 | 0.00253<br>(0.00152 -<br>0.00353) | 0.001<br>0.0049 |
| **ELOVL2**<br>**(DPA/EPA)** | 5.74<br>[4.67 -<br>6.94]*** | 5.53<br>[4.47 -<br>6.53]*** | 5.18<br>[4.04 -<br>6.17]** | 4.92<br>[3.96 -<br>6.00] | 4.85<br>[3.74 -<br>5.86] | < 0.001 | -0.202 (-<br>0.228 - -<br>0.173) | < 0.001 | -0.02228 (-<br>0.02519 - -<br>0.01936) | < 0.001<br>0.0441 |
| **ELOVL5**<br>**(DGLA/GLA)** | 32.7<br>[26.3 -<br>41.3]*** | 31.1<br>[24.4 -<br>38.7]*** | 28.6<br>[23.0 -<br>35.9] | 27.1<br>[21.9 -<br>33.7] | 27.4<br>[22.0 - 34.8] | < 0.001 | -0.177 (-<br>0.205 - -<br>0.149) | < 0.001 | -0.1217<br>(-0.1400 -<br>-0.1034) | < 0.001<br>0.0339 |
| **ELOVL6**<br>**(SA/PA)** | 0.69<br>[0.65 -<br>0.72] | 0.69<br>[0.65 -<br>0.72] | 0.69<br>[0.65 -<br>0.72] | 0.69<br>[0.66 -<br>0.73] | 0.69<br>[0.66 -<br>0.72] | 0.438 | 0.013 (-<br>0.016 -<br>0.041) | 0.372 | 7.097<br>e-005<br>(-2.067<br>e-005 -<br>0.000162) | 0.129<br>0.0005 |
| **SCD16**<br>**(POA/PA)** | 0.013<br>[0.010 -<br>0.015]*** | 0.013<br>[0.011 -<br>0.016] | 0.014<br>[0.011 -<br>0.018]** | 0.014<br>[0.011 -<br>0.018] | 0.013<br>[0.011 -<br>0.017] | < 0.001 | 0.072<br>(0.041 -<br>0.099) | < 0.001 | 2.941<br>e-005<br>(1.895<br>e-005 -<br>3.987<br>e-005) | < 0.001<br>0.0062 |
| **SCD18**<br>**(OA/SA)** | 0.87<br>[0.76 -<br>0.89]*** | 0.84<br>[0.77 -<br>0.91]*** | 0.84<br>[0.78 -<br>0.91]*** | 0.85<br>[0.78 -<br>0.92]** | 0.86<br>[0.79 -<br>0.93] | < 0.001 | 0.139<br>(0.110 -<br>0.167) | < 0.001 | 0.000917<br>(0.000734<br>- 0.00110) | < 0.001<br>0.0195 |

Values are presented as median [interquartile range, Q1-Q3]. Ratio-based indices are proxies of enzyme activity and do not represent direct enzymatic measurements. Overall p values denote differences across the five age
groups (Kruskal-Wallis test); post hoc comparisons versus the 65-79-year reference group were performed using Dunn's multiple comparisons test with multiplicity-adjusted p values, and asterisks indicate significant differences versus 65-79 years (\*p < 0.05, \*\*p < 0.01, \*\*\*p < 0.001; adjusted). Associations with age as a continuous variable are shown as Spearman rank correlation coefficients ( $\rho$ ) with 95% confidence intervals (CI) and two-sided p values. Linear regression slopes (per 1-year increase in age) are given with 95% CI, two-sided p values, and $R^2$ . Table layout and statistical approach adapted from Yu et al.

D5D (AA/DGLA) and the composite n-6 proxy AA/LA were positively associated with age, both as a group effect (both overall p < 0.001) and as continuous associations (ρ = 0.122 and ρ = 0.243, respectively; both p < 0.001). The D6D proxy based on γ-linolenic acid (GLA)/LA was also positively associated with age (ρ = 0.225, p < 0.001), which is a stronger and more consistent signal than in the clinical cohort, where D6D associations were weak and dependent on the product-to-precursor pair used. The more distal D6D proxy DGLA/LA showed a much weaker positive association (ρ = 0.070).

Elongation-related proxies were predominantly inversely associated with age: ELOVL2 (docosatetraenoic acid (DTA)/AA) ρ = ™0.188, ELOVL2 (docosapentaenoic acid (DPA)/EPA) ρ = ™0.201 and ELOVL5 (DGLA/GLA) ρ = ™0.177 (all p < 0.001). The exception was ELOVL2 (DHA/DPA), which was positively associated with age (ρ = 0.101, p < 0.001).

ELOVL6 (stearic acid (SA)/palmitic acid (PA)) showed no association with age in either the group comparison (p = 0.438) or the continuous analysis (ρ = 0.013, p = 0.372); this differs from the Brandenburg cohort, where ELOVL6 was inversely associated with age. Among the SCD indices, SCD18 (oleic acid (OA)/SA) was positively associated with age (ρ = 0.139, p < 0.001) and SCD16 (palmitoleic acid (POA)/PA) weakly so (ρ = 0.070, p < 0.001) without a monotone gradient across strata. Across all indices, age explained at most 6 % of the variance (AA/LA, R^2^ = 0.061).

### 3.6 Omega-3 index and PhenoAge acceleration

In the fasting subsample with biochemical measurements including fasting glucose measurements plus the other parameters necessary for PhenoAge calculation (n = 2822; median age 56.0 [39 - 66] years), median PhenoAge was 56.0 [38.4 - 67.1] years and median PhenoAgeAccel was ™1.4 [-5.0 - 3.0] years. Treating PhenoAgeAccel as a continuous variable demonstrates negative correlations of long-chain n-3 PUFA with PhenoAgeAccel (Figure 3). EPA + DHA was positively correlated with age (ρ = 0.18, p < 0.001; slope 0.017 percentage points per year, R^2^ = 0.030), as were EPA (ρ = 0.21, p < 0.001) and DHA (ρ = 0.17, p < 0.001).

**Figure 3.**
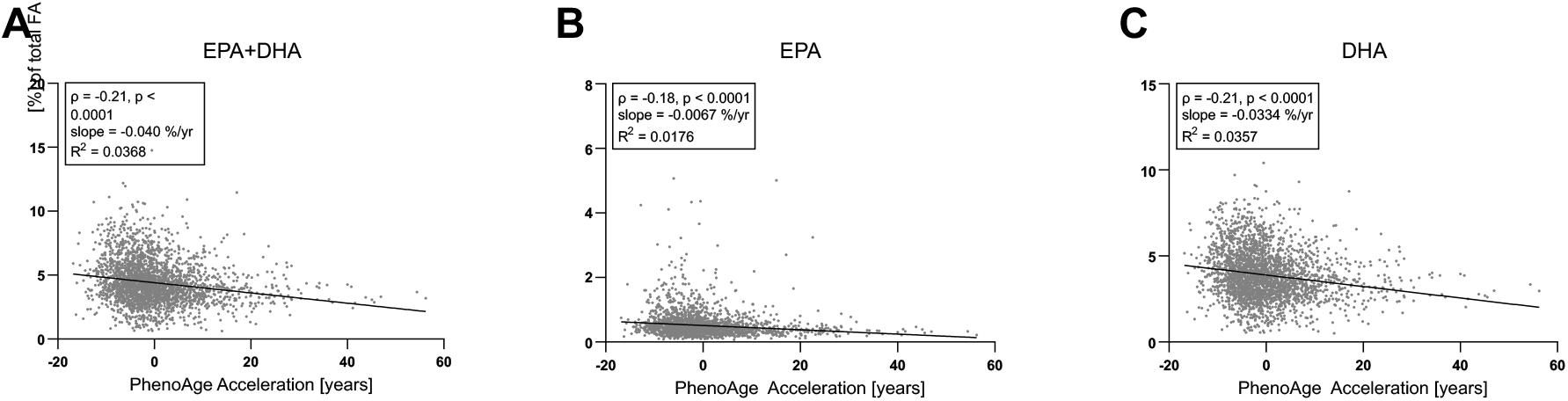
Associations of erythrocyte n-3 PUFA with PhenoAge acceleration. (A) Scatter plot of erythrocyte EPA+DHA (% of total fatty acids; y-axis) against PhenoAge acceleration (PhenoAgeAccel) (x-axis) for the full cohort (n = 2822). The solid line represents the best-fit simple linear regression with; correlation coefficients, regression slopes, p and R2 are shown. PhenoAgeAccel was defined as the residual from the linear regression of PhenoAge on chronological age; positive values indicate biological aging accelerated relative to chronological age. Corresponding scatter plots of the individual n-3 PUFAs are shown in (B) for erythrocyte EPA and (C) for DHA.

Using the conventional omega-3 index categories, adjusted PhenoAge acceleration relative to an index below 4 % was ™1.7 years for 4-< 6 %, ™2,7 years for 6-< 8 % and ™4.7 years for ≥ 8 % (all p < 0.001) (Table 4 and Figure 4).

**Table 4.**
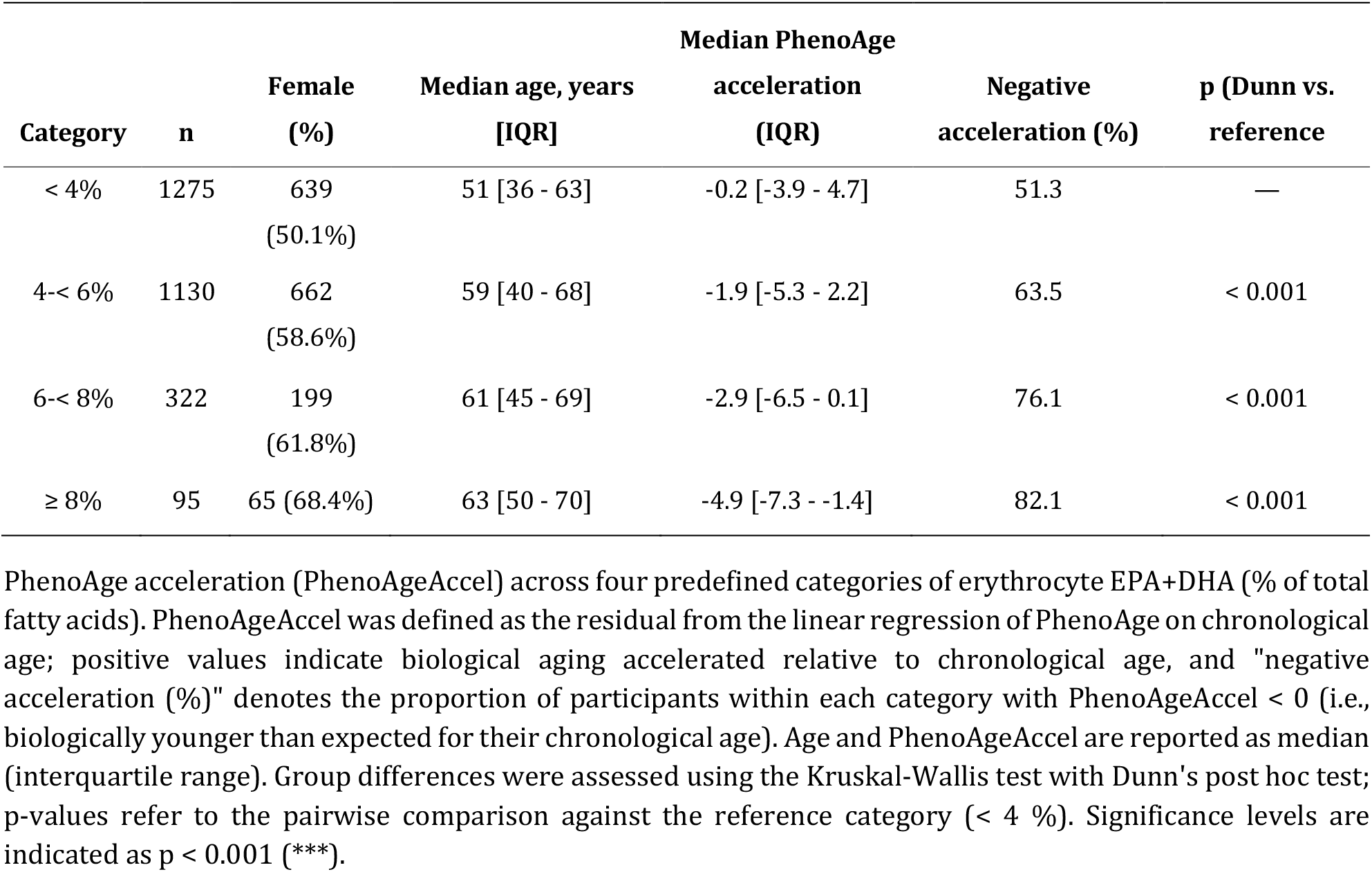
Erythrocyte EPA+DHA categories and PhenoAge acceleration.

**Figure 4.**
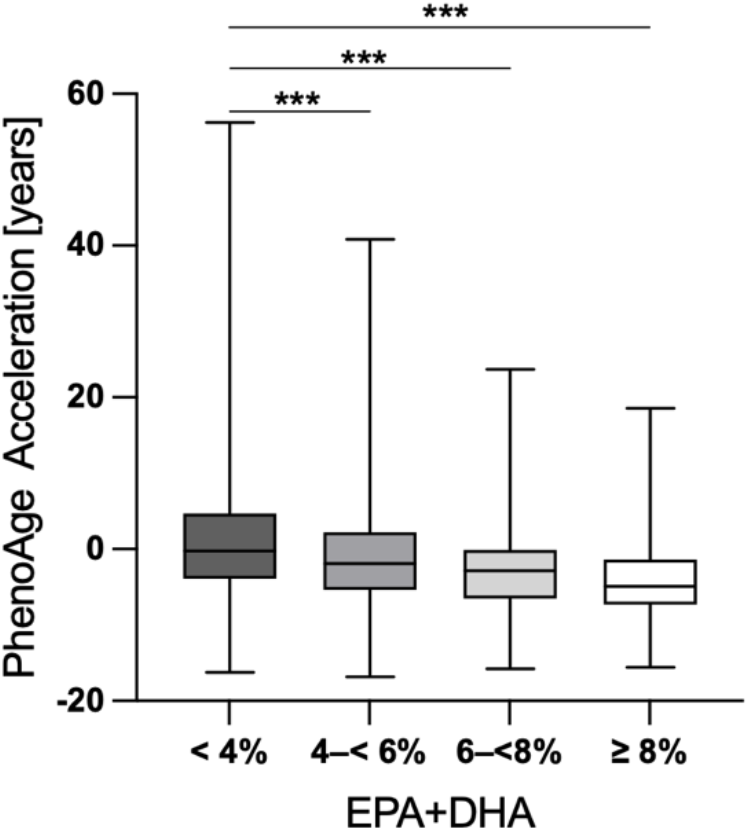
Erythrocyte EPA+DHA and PhenoAge acceleration. PhenoAge acceleration (PhenoAgeAccel) across predefined categories of erythrocyte EPA+DHA. PhenoAgeAccel (y-axis) is shown for four omega-3 Index categories (% of total fatty acids): Group 1 = < 4 % (n = 1275), Group 2 = 4-< 6 % (n = 1130), Group 3 = 6-< 8 % (n = 322), and Group 4 = ≥ 8 % (n = 95). Boxes show the median and interquartile range (25th-75th percentiles); whiskers extend from minimum to maximum. Group differences were assessed using the Kruskal-Wallis test with Dunn’s post hoc test, with Group 1 (< 4 %) as the reference group. Significance levels are indicated as p < 0.001 (***).

We also performed an analysis comparing n-3 PUFA levels across tertiles of PhenoAgeAccel. In the tertile with the lowest PhenoAgeAccel, i.e. those phenotypically youngest by this proxy measurement (with a median PhenoAgeAccel of -6.4 years), levels of EPA+DHA, as well as both individual n-3 PUFA were significantly higher as compared to those in the phenotypically oldest tertile (with a median PhenoAgeAccel of 5.7 years). This was also true for EPA and DHA as well as EPA+DHA levels compared between the middle tertile (with a median PhenoAgeAccel of -1.4 years) , demonstrating that higher EPA+DHA levels are associated with lower PhenoAgeAccel throughout (Table 5).

**Table 5.** Participant characteristics and erythrocyte n-3 fatty acids across tertiles of PhenoAge acceleration.

| Tertile<br>PhenoAge<br>Acceleration<br>(n) | Female<br>(%) | Median Age<br>[IQR]<br>(years) | Median PhenoAge<br>[IQR]<br>(years) | Median PhenoAge<br>Acceleration<br>[IQR]<br>(years) | EPA+DHA<br>[%] | EPA<br>[%] | DHA<br>[%] |
| --- | --- | --- | --- | --- | --- | --- | --- |
| T1 (n = 940) | 603<br>(64.1%) | 58 [41-67] | 52 [35 - 61] | -6.4 [-8.2 - -5.0] | 4.5<br>[3.6 - 5.8]*** | 0.46<br>[0.35 - 0.65]*** | 4.0<br>[3.2 - 5.1]*** |
| T2 (n = 941) | 438<br>(46.6%) | 53 [36-65] | 53 [35 - 65] | -1.4 [-2.5 - -0.2] | 4.2<br>[3.3 - 5.2]*** | 0.42<br>[0.33 - 0.56]*** | 3.7<br>[2.9 - 4.7]*** |
| T3 (n = 940) | 479<br>(50.9%) | 56 [40-66] | 65 [47 - 76] | 5.7 [3.0 - 10.4] | 3.9<br>[3.1 - 4.7] | 0.39<br>[0.31 - 0.51] | 3.4<br>[2.8 - 4.2] |
| Overall p | < 0.001 | < 0.01 | n.d. | n.d. | < 0.001 | < 0.001 | < 0.001 |

Characteristics of the study population (n = 2822) stratified by tertiles (T1 - T3) of PhenoAge acceleration (PhenoAgeAccel). PhenoAgeAccel was defined as the residual from the regression of Phenotypic Age on chronological age; positive values indicate biological aging accelerated relative to chronological age. Tertiles correspond to increasing biological aging: T1 = decelerated/biologically youngest, T2 = average, and T3 = accelerated/biologically oldest. For each tertile, the number of female, and median age, Phenotypic Age, PhenoAge acceleration, and erythrocyte EPA+DHA, EPA (C20:5n-3), and DHA (C22:6n-3), each expressed as % of total fatty acids, are shown. Continuous variables are reported as median [interquartile range, IQR]. Overall p-values were determined using the Kruskal-Wallis test, with Dunn's post hoc test and T3 (accelerated) as the reference group. For sex distribution the Chi-square test was used. Overall p-values were not determined (n.d.) for Phenotypic Age and PhenoAgeAccel, as these variables define or are structurally related to the tertile grouping.

## 4 Discussion

This study uses data from the NHANES cycle 2021-2023 in an attempt to further address n-3 PUFA blood levels with regard to age and aging. Previous analyses have established increasing n-3 PUFA levels with age, as well as associations of increased n-3 PUFA with healthy ageing decreased premature mortality, as described above.

In a nationally representative sample of 4863 US adults, we reproduced the principal age-related patterns in erythrocyte fatty acid composition and in desaturase and elongase proxy indices that we had recently described in a clinical cohort from Brandenburg, Germany. Furthermore, we found that a higher omega-3 index was consistently associated with lower PhenoAgeAccel, an established marker of biological as compared to chronological aging.

When regarding associations between chronological age and PUFA, total n-3 PUFA and EPA + DHA were higher, and total n-6 PUFA lower, in older participants. Second, EPA and DHA increased with age while LA and DGLA decreased. Third, AA was comparatively stable across age strata. Fourth, D5D and AA/LA rose with age while ELOVL2-related elongation proxies (DTA/AA, DPA/EPA) and ELOVL5 fell. These findings in the NHANES cohort are consistent with our observations in a metabolic patient cohort from Germany (Yu et al., 2026).This replication of patterns across two populations of different composition, in different countries, and with independent analytical platforms strengthens the case that these are genuine age-related shifts in PUFA handling rather than artefacts of one clinical case-mix and environment.

In this study we also assessed associations of EPA + DHA, analogous to the omega-3 index, with biomarker-based PhenoAge, which has been established as a marker of biological age as compared to chronological age in data from a previous NHANES cycle (Levine et al., 2018; Liu et al., 2018). The association we observed — approximately 4.7 fewer years of PhenoAgeAccel between the group with an omega-3 index below 4 % as compared to one above 8 % — is substantially larger than the effect of n-3 supplementation on epigenetic aging measured in a randomized trial, where 1 g/day for three years slowed biological clocks by roughly 3 to 4 months (Bischoff-Ferrari et al., 2025).

Long-chain n-3 PUFA derive predominantly from dietary EPA and DHA, with less than 10 % contributed by endogenous conversion from ALA (Burdge and Calder, 2005; Brenna et al., 2009), therefore, higher EPA and DHA in older adults might reflect other effects such as dietary intake and increased supplementation of EPA and DHA than in younger probands. Beyond diet, further factors including behaviors like smoking, physical activity, medication use, and genetic variation, also influence PUFA status and cannot be excluded as contributors to the observed associations (Hodson et al., 2008).

The previously published evaluation of PUFA measurements in this NHANES cycle showed that supplement users had a higher omega-3 index (Powers et al., 2026). Furthermore, a higher omega-3 index was associated with not smoking, a healthy weight, higher income and higher education (Powers et al., 2026). In our analyses shown here, neither diet, smoking, physical activity, medications, genetics nor supplement use was included.

Biomarker based PhenoAge has been established in recent years as a tool to evaluate biological age in a variety of contexts, and it has been shown to be a prognostic marker in some studies, e.g. (Shao et al., 2026). In our evaluation presented here, we found a clear negative correlation of PhenoAgeAccel with increasing levels of EPA and DHA, indicating that higher n-3 PUFA might contribute to lowering biological age. This was found throughout the whole spectrum of EPA and DHA levels - even moderately higher n-3 PUFA blood content was already associated with significant decreases of PhenoAge acceleration. These observations are consistent with previous data implicating n-3 PUFA to reduce mortality risk.

### Limitations

This analysis is cross-sectional and cannot support causal inference. The ratio-based indices are proxies of pathway balance, not measurements of enzyme activity, and are influenced by diet, adiposity, inflammation and fatty acid desaturase (*FADS)* genotype (Lattka et al., 2010; Gonzalez-Soto and Mutch, 2021; Loukil et al., 2024; Rabehl et al., 2024). No genetic data were used here.

The PhenoAge analysis was restricted to the fasting subsample (n = 2822 of 4863), which is not a random subset of the full sample. Furthermore, chronological age is only given in full years, decreasing the accuracy of the calculation. The descriptive fatty acid analyses are unweighted and should not be read as national prevalence estimates. Weighted national estimates for this cycle are reported by Powers et al. (Powers et al., 2026).

No mortality linkage is available for this NHANES cycle, so PhenoAgeAccel could not be validated against outcomes within these data.

In addition, PhenoAge is calculated from inflammatory, hematological and metabolic markers, several of which could plausibly alter fatty acid handling, incorporation into erythrocyte membranes or erythrocyte turnover. Chronic inflammation in particular affects both PhenoAge components and PUFA metabolism, and our analyses cannot separate these effects.

Diet, smoking, physical activity and n-3 supplement use were not included in our models and are variables that could modify the interpretation.

### Conclusion

Age-related shifts in erythrocyte PUFA composition and in desaturase and elongase proxy indices previously described in are reproduced in a nationally representative US population, supporting their generality while leaving their mechanistic basis open. Furthermore, in this population a higher omega-3 index was associated with lower phenotypic age acceleration, supporting a role of long-chain n-3 PUFA status to slow biological aging. Longitudinal and interventional designs with dietary and supplement information will be necessary to establish causal effects.

## Data Availability

All data produced in the present study are available upon reasonable request to the authors

## Statements

### Data availability statement

All data analyzed in this study are publicly available from the NHANES repository of the Centers for Disease Control and Prevention (https://wwwn.cdc.gov/nchs/nhanes/), cycle August 2021-August 2023. Analysis code is available from the corresponding author on request.

### Ethics statement

NHANES is approved by the NCHS Research Ethics Review Board and all participants provided written informed consent. The present analysis used publicly available deidentified data and did not require additional ethical approval.

### Author contributions

AP: Investigation, Formal Analysis, Visualization, Writing - review and editing, NR: Methodology, Data curation Investigation, Analysis, Writing - review and editing, KHW: Conceptualization, Investigation, Methodology, Resources, Writing - original draft, review and editing.

### Generative AI statement

The authors declare that generative AI was used in the creation of this manuscript. During the preparation of this work the authors used Gemini 3 Flash and Opus 5.0 to improve langue and help with drafting, as well as for literature and methodology searches and plausibility checking. After using these tools/service, the authors reviewed, selected corrected, modified and edited the information as needed and take full responsibility for the content of the submitted manuscript.

